# Can ocular dominance plasticity provide a general index to visual plasticity to personalize treatment in amblyopia?

**DOI:** 10.1101/2020.03.26.20044701

**Authors:** Chunwen Tao, Zhifen He, Yiya Chen, Jiawei Zhou, Robert F. Hess

**Affiliations:** School of Ophthalmology and Optometry and Eye hospital, and State Key Laboratory of Ophthalmology, Optometry and Vision Science, Wenzhou Medical University, Wenzhou, Zhejiang, PR China; McGill Vision Research, Dept. Ophthalmology and Vision Sciences, McGill University, Montreal PQ, Canada

## Abstract

**Purpose:** Recently, Lunghi et al showed that amblyopic eye’s visual acuity per se after 2 months of occlusion therapy could be predicted by a homeostatic plasticity, i.e., the temporary shift of ocular dominance observed after a 2-hour monocular deprivation, in children with anisometropic amblyopia(Lunghi et al., 2016). In this study, we assess whether the visual acuity *improvement* of the amblyopic eye measured after 2 months of occlusion therapy could be predicted by this plasticity.

**Methods:** Seven children (6.86 ± 1.46 years old; SD) with anisometropic amblyopia participated in this study. All patients were newly diagnosed and had no treatment history before participating in our study. They had finished 2 months of refractive adaptation and then received a 4-hour daily fellow eye patching therapy with an opaque patch for a 2-month period. Best-corrected visual acuity of the amblyopic eye was measured before and after the patching therapy. The homeostatic plasticity was assessed by measuring the temporary shift of ocular dominance observed after 2 hours of occlusion for the amblyopic eye before the treatment started. A binocular phase combination paradigm was used for this test.

**Results:** We found that there was no significant correlation between the temporary shift of ocular dominance observed after 2 hours of occlusion for the amblyopic eye before the treatment started and the visual acuity gain obtained by the amblyopic eye from 2-month of classical patching therapy. This result involving the short-term patching of the amblyopic eye is consistent with a reanalysis of Lunghi et al’ s data.

**Conclusions:** Ocular dominance plasticity does not provide an index of cortical plasticity in the general sense such that it could be used to predict acuity outcomes from longer term classical patching.

## Introduction

There is a considerable variability in the response to amblyopic treatment, be it classical occlusion therapy(Stewart et al., 2005) or binocular therapy(Hess and Thompson, 2015) across a population of amblyopes of all ages. Not all of this variance can be attributed to differences in compliance(Stewart et al., 2005; Stewart et al., 2007b; Fronius et al., 2014), leading us to the inescapable conclusion that some amblyopes have brains that are more capable of change, in other words, more plastic, than others. Unfortunately, there is no way of knowing which patients are more likely to respond to the treatment before the therapy begins, in the main (though see,(Stewart et al., 2007a)) it is not until the end that the responders can be separated from the non-responders. If we had some general measure of visual cortical plasticity it may be possible to personalize the present treatment and avoid subjecting patients who are unlikely to respond to months or years of the sort of disruption that patching the one good eye produces.

Ocular dominance plasticity is a recently observed phenomenon(Lunghi et al., 2011), (Lunghi et al., 2011) in which the ocular dominance of normal adults can be manipulated by short-term visual deprivation of one eye. Comparable effects occur in adults with amblyopia(Zhou et al., 2013c). This has been extensively studied using psychophysics(Lunghi et al., 2011; Lunghi et al., 2013; Zhou et al., 2013a; Zhou et al., 2013c; Zhou et al., 2014) (Lunghi and Sale, 2015; Zhou and Hess, 2016; Zhou et al., 2017a; Zhou et al., 2017b), electrophysiology (Lunghi et al., 2015a; Zhou et al., 2015) and brain imaging (Lunghi et al., 2015b; Chadnova et al., 2017; Binda et al., 2018). There is evidence that it involves a reciprocal change in sensitivity of each eye’s input; the previously patched eye becomes more dominant and the previously unpatched eye becomes less dominant, i.e., a homeostatic form of plasticity (Zhou et al., 2013a; Chadnova et al., 2017). It represents a measure of visual cortical plasticity and it could potentially represent a general index to how modifiable the visual areas of the brain are and as such could provide a measure that might allow greater predictability for more long-term procedures such as amblyopia patching therapy which is currently associated with a high level of variability in terms of its effectiveness across a population of amblyopes. If this was the case, patching therapy could be personalized using the measure, ocular dominance plasticity in terms of hours per day and total duration. One study has addressed this very interesting question using the measure, short-term ocular dominance plasticity, and concluded that children who exhibit a higher degree of short-term ocular dominance plasticity (determined after a 2-hour period of patching session) go on to obtain larger amblyopic eye “recovery rate” after a long-term (months) patching procedures (Lunghi et al., 2016). It should be noted that, the amblyopic eye “recovery rate” was defined as the absolute final visual acuity after the long-term patching in Lunghi et al’s study (Lunghi et al., 2016). It would be interesting to further investigate whether large changes in ocular dominance plasticity were able to predict large *improvements* (i.e., difference between initial acuity before occlusion therapy and that found after occlusion therapy) in visual acuity (i.e., the effects of patching therapy) as a result of long-term patching.

We directly tested this idea in this study. Initially we measured ocular dominance plasticity by patching the amblyopic eye for 2 hours. Second, classical occlusion therapy with an opaque patch occluding the fellow eye (4hrs/day for 2 months) was undertaken in 7 newly diagnosed patients. Any patient who needed an update to the spectacle correction was allowed a 2-month period before undertaking the experiment.

## Methods

### Participants

Seven children (6.86 ± 1.46 years old; SD) who had anisometropic or ametropic amblyopia and were able to perform the binocular phase combination task after practice participated in this study. All patients were newly diagnosed and had no treatment history before participated in our study. The clinical details of the patients and their visual acuity before and after 2 months of treatment are provided in Table 1. All participates were naive to the purpose of the study. Written informed consent was obtained from their parents or guardians before the start of the experiment. This study followed the tenets of the Declaration of Helsinki and was approved by the Ethics Committee of Wenzhou Medical University and McGill University.

**Table 1.** Visual acuity before and after 2 months of treatment.

| ID | Age | Cycloplegic | Visual acuity (logMAR) |  |  |  | Balance point |
| --- | --- | --- | --- | --- | --- | --- | --- |
|  |  | refractive errors | Before treatment |  | 2 months of treatment |  | (AE/FE) |
|  |  | (OD/OS) | OD | OS | OD | OS | before treatment |
| S1 | 5 | - 1.00/ - 1.00 ×180<br>- 6.00/ - 2.00 ×180 | 0.275 | 0.575 | 0.175 | 0.375 | 0.432 |
| S2 | 6 | + 1.50<br>+ 5.00 | -0.025 | 0.575 | -0.025 | 0.275 | 0.285 |
| S3 | 8 | Plano<br>+ 2.50/ +1.75 × 80 | -0.025 | 0.575 | -0.025 | 0.475 | 0.192 |
| S4 | 6 | + 3.50<br>+ 4.00/ + 0.75 × 95 | 0.175 | 0.675 | 0.175 | 0.275 | 0.158 |
| S5 | 8 | + 4.50<br>Plano | 0.475 | -0.025 | 0.275 | -0.025 | 0.329 |
| S6 | 9 | + 4.00<br>Plano | 0.575 | -0.125 | 0.4 | -0.125 | 0.135 |
| S7 | 6 | Plano<br>+ 2.00/ + 1.75 × 85 | -0.025 | 0.875 | -0.025 | 0.775 | 0.234 |

### Apparatus

The stimuli for the short-term monocular deprivation measurement were generated and controlled by a PC computer running Matlab (MathWorks, Natick, MA) with PsychTool Box 3.0.9 extension (Brainard, 1997; Pelli, 1997). The stimuli were presented on a gamma-corrected LG D2342PY 3D LED screen (LG Life Science, Korea) with a 1920 × 1080 resolution and a 60 Hz refresh rate. Subjects viewed the display dichoptically with polarized glasses in a dimly lit room at a viewing distance of 136 cm. The background luminance was 46.2 cd/m^2^ on the screen and 18.8 cd/m^2^ through the polarized glasses. Patients’ best-corrected visual acuity was measured monocularly using the Logarithmic Tumbling E Chart (Mou, 1966) at 5 m.

### Design

In this study, the treatment effect of 2 months patching therapy (4-hour daily fellow eye patching with an opaque patch) was tested after a 2-month of refractive adaptation. The short-term monocular deprivation effect was quantified in an initial experiment by the shift of ocular dominance in binocular phase combination after 2-hour of amblyopic eye patching(Zhou et al., 2013c). An illustration of the experimental design is provided in Figure 1.

**Figure 1.**
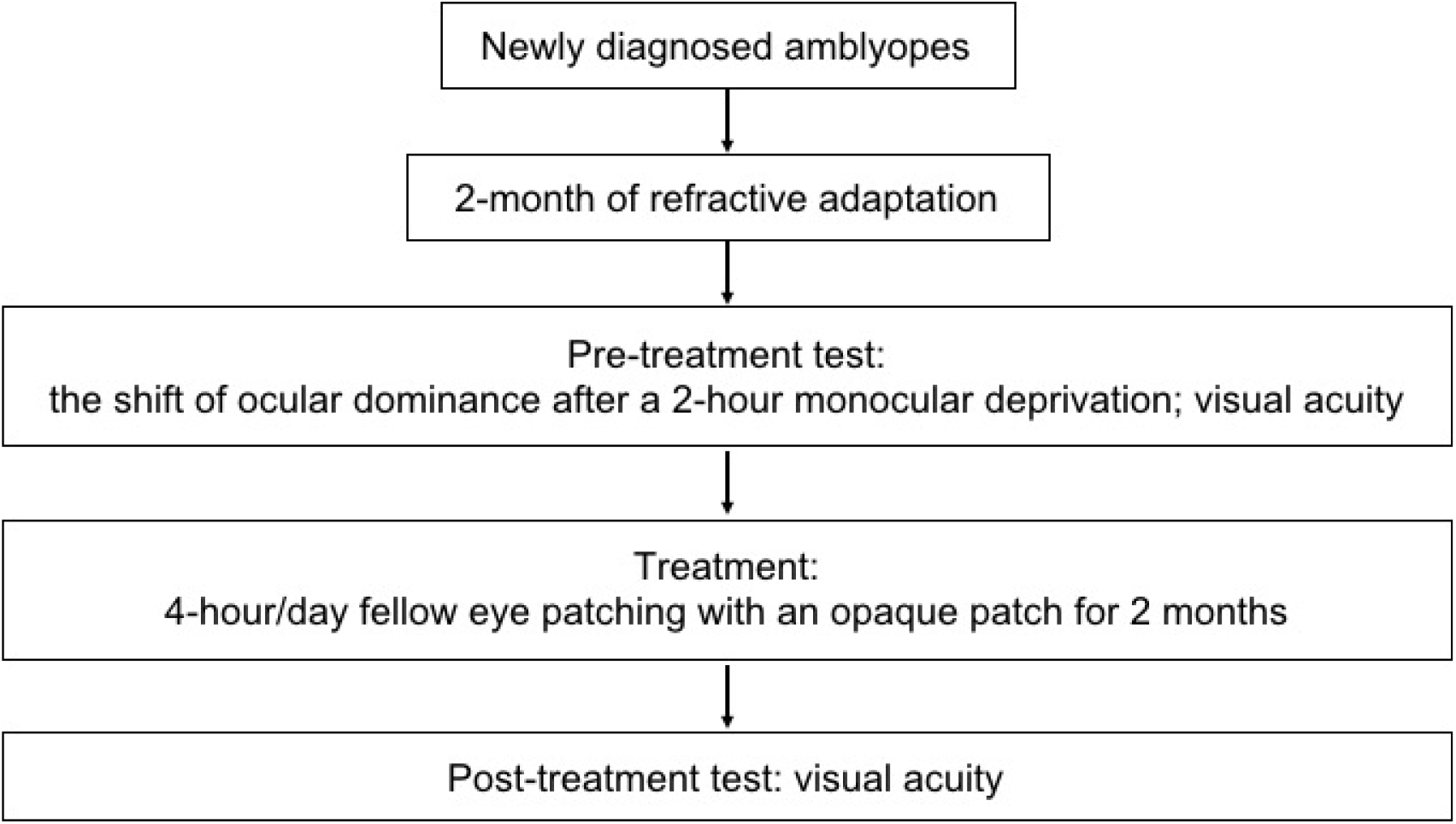
An illustration of the experimental design. Seven newly diagnosed child amblyopes (6.86 ± 1.46 years old; SD) participated, in which the treatment effect of 2 months patching therapy (4-hour daily patching with an opaque patch) was tested after a 2-month of refractive adaptation. The short-term monocular deprivation effect was quantified by the shift of ocular dominance in binocular phase combination after 2-hour of amblyopic eye patching before the initial of the treatment.

### Procedure and stimuli

Similar to our previous studies (Zhou et al., 2013a), the short-term monocular deprivation effect was tested with a binocular phase combination task. In the measure, two horizontal sine-wave gratings (1 cycle/°, 2° × 2°), with equal and opposite phase shifts (+22.5° and - 22.5°) relative to the center of the screen were dichoptically presented to the two eyes. The perceived phase of fused stimuli was 0° when the two eyes contributed equally to binocular fusion. The interocular contrast ratio at that condition was the balance point in binocular phase combination. We firstly tested this balance point for each patient with the contrast of the stimuli in the amblyopic eye set as 100%. This was achieved by measuring individual’s binocularly perceived phase at interocular contrast ratios of 0, 0.1, 0.2, 0.4, 0.8 and 1; and the binocularly perceived phase vs. interocular contrast ratio (PvR) curve was fitted with a contrast-gain control model(Ding and Sperling, 2006; Zhou et al., 2013b). One to three hours of practice trials were provided before we conducted of the main study to make sure patients understood the task and had a reliable performance in the binocular phase combination task. Individuals’ PvR curves measured before treatment are provided in Figure 1. The balance points of patients before treatment are provided in Table 1.

We then fixed the contrast of the stimuli in the two eyes based on individual’s balance point and tested individuals’ binocularly perceived phase before and after a 2-hour of monocular deprivation of the amblyopic eye with an opaque patch. Two stimuli configurations were used for measuring the binocularly perceived phase to account for positional bias: +22.5° phase in the amblyopic eye and -22.5° phase in the fellow eye and -22.5° phase in the amblyopic eye and +22.5° phase in the fellow eye. The half of the difference between these two configurations was calculated as the binocularly perceived phase. Each session of binocularly perceived phase measurement contains 16 trails (2 configurations * 8 repetitions). The two configurations were randomly assigned in different trials. In each trial, observers were asked to adjust the position of a flanking reference line to locate the middle of the dark strip of the binocularly perceived grating to indicate its phase. A high-contrast frame (0.11° in width and 6° in length) with four white diagonal lines (0.11° in width and 2.83° in length) was continually presented surrounding the grating in each eye to help observers maintain fusion. Subjects normally needed around 3 minutes to finish one measurement session. We tested 3 sessions of binocularly perceived phase within 10 minutes after patients finished the 2-hour of monocular deprivation. We averaged the results of these 3 sessions and then calculated the difference between the average post-patching perceived phase and the baseline to get the ocular dominance difference index after the 2-hour of monocular deprivation. We also normalized individuals’ ocular dominance difference index to the largest one in this group. This normalization ensured that the normalized ocular dominance difference index ranged from - 0.5 to 1, similar to the range reported in Lunghi et al’s (Lunghi et al., 2016). The normalization itself would not change any correlation analysis we conducted in this study.

For the best-corrected visual acuity measure, we asked patients to read the optotypes one after another and stopped when they could not respond within 10 s. We calculated their percentage correct at different lines of the Logarithmic visual acuity Chart. We then used linear interpolation to calculate the score associated with 75% correct judgments. This score was defined as patients’ visual acuity.

## Results

In Figure 2, we plot individual’s binocularly perceived phase as a function of the interocular contrast ratio measured before the treatment. The contrast-gain control model fits well to the data, with an average goodness-of-fit of 0.941 ± 0.054 (mean ± SD). This is similar to our previous observation(Zhou et al., 2013b) in adults with amblyopia (0.951 ± 0.022), indicating that our patients in this study were able to make reliable measurements with the binocular phase combination task before the treatment. Also, similar to our previous observation(Zhou et al., 2013b), there was a trend toward decreasing contrast ratios being associated with increasing interocular visual acuity differences (r = -0.59, *P* = 0.16).

**Figure 2.**
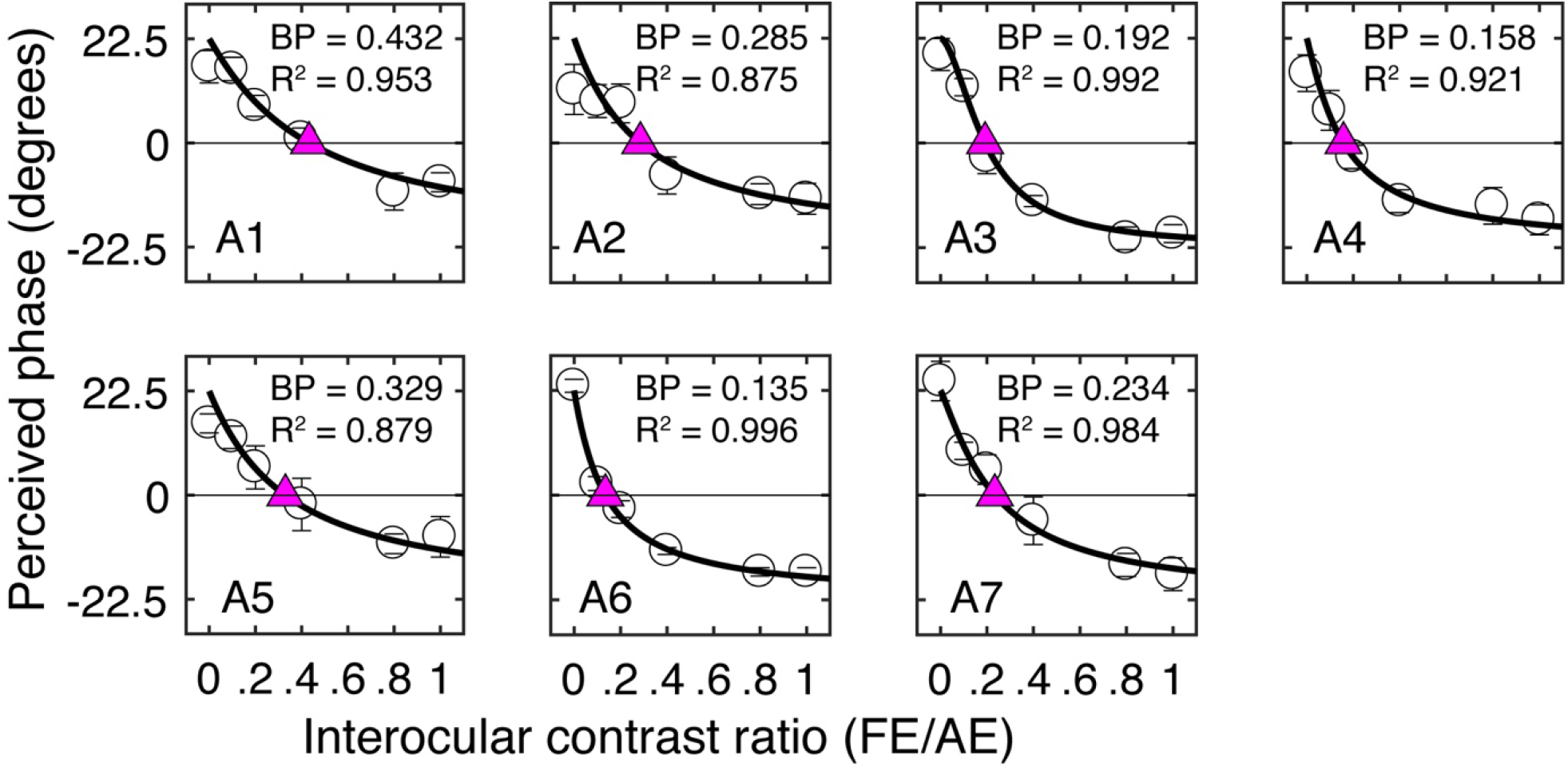
Individual’s binocularly perceived phase as a function of the interocular contrast ratio measured before the patching treatment. Each panel plots results of one patient. Error bars represent standard errors from 8 repetitions of the test. The curve in each panel represents fits with a contrast-gain-control model(Zhou et al., 2013b). The purple triangle represents where the two eyes were balanced. The corresponding interocular contrast ratio (in short “BP”) and the goodness-of-fit are provided in each panel.

Long term patching therapy (4hr/day for 2 months) significantly improved the visual acuity of the amblyopic eye in our patients, from an average of 0.62 ± 0.05 to 0.41 ± 0.07 (logMAR): Z = -2.38, *P* = 0.018, 2-tailed Wilcoxon Signed Ranks Test. In Figure 3A, we plot the amblyopic eye acuity improvement after 2 months of treatment as a function of the normalized short-term ocular dominance index difference for the seven patients in our study. The normalized ocular dominance index difference was larger than 0 in five of the seven patients, which indicates a strengthening of the patched eye after the 2-hour short-term monocular deprivation. Two of the seven patients had a shift of ocular dominance in the reversal direction. This pattern of result was similar to that previously reported by Lunghi et al (2016), in which one of their 10 patients had a shift of ocular dominance in the reversal way (see Figure 3B). A 2-tailed Pearson correlation analysis showed that the correlation between the amblyopic eye acuity improvement after 2 months of treatment and the normalized ocular dominance index difference after 2 hrs of amblyopic patching was not significant: r = 0.20, *P* = 0.66.

**Figure 3.**
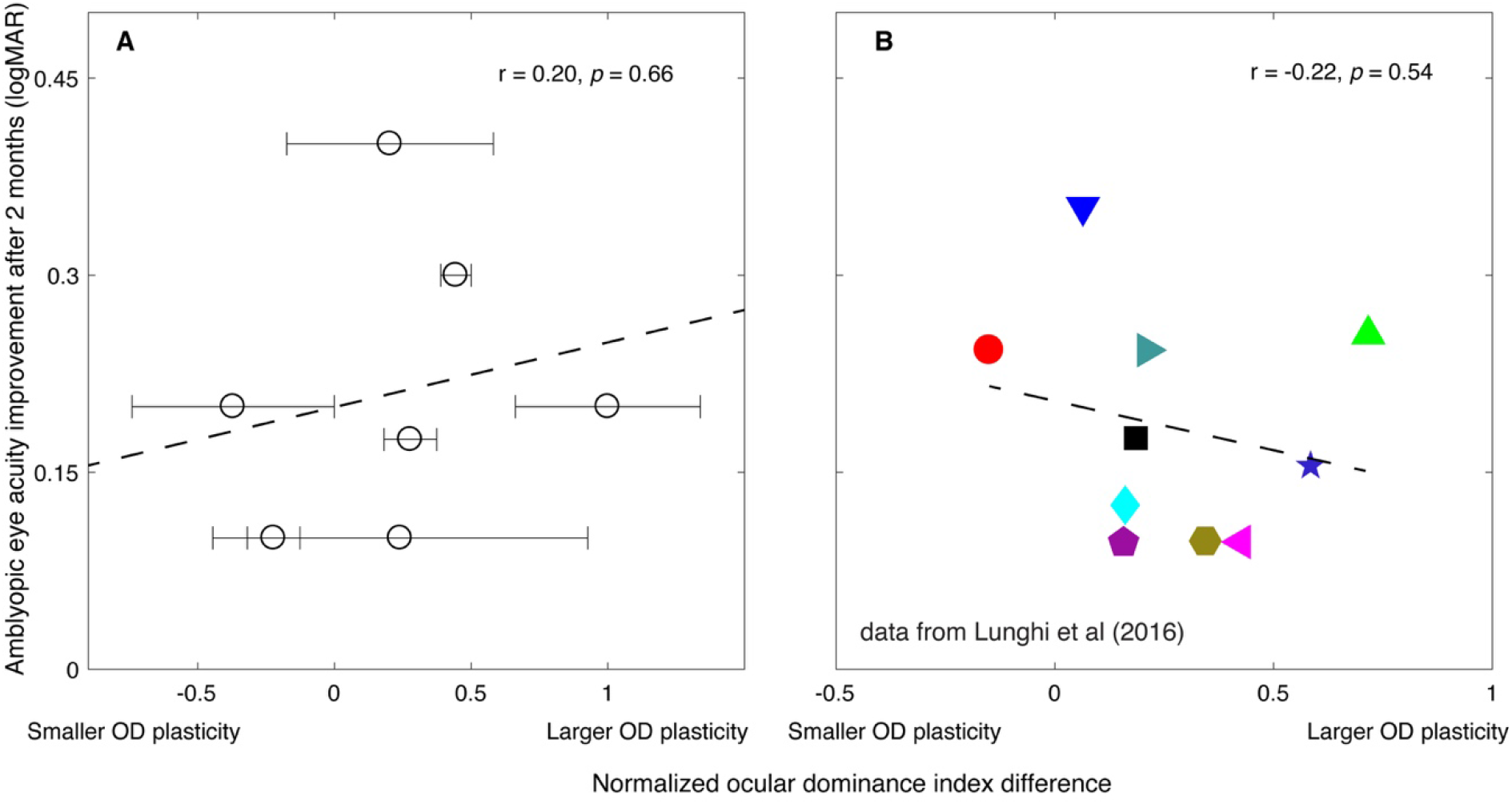
The relationship between the amblyopic eye acuity improvement after 2 months of treatment and the short-term monocular deprivation effect. (A). Results from the present study. The horizontal axis represents the effect of short-term monocular deprivation on the ocular dominance (OD) shift. The value, if larger than 0, indicates the patched eye was getting stronger after the short-term monocular deprivation. The larger the value indicates the larger OD plasticity. The dashed line is a linear fit of the data. The error bars represent standard deviation based on the three post-test sessions measured within 10 minutes after patients finished the 2-hour of monocular deprivation. A 2-tailed Pearson correlation analysis showed that the correlation between the amblyopic eye acuity improvement after 2 months of treatment and the normalized ocular dominance index difference was not significant: r = 0.20, *P* = 0.66. (B). Reports from Lunghi et al’s study(Lunghi et al., 2016). Their work is licensed under a Creative Commons Attribution-NonCommercial-NoDerivatives 4.0 International License. The results from Figure 4B and Table 1 in Lunghi et al (2016)’s paper are replotted here. Ten patients (6.2 ± 1.0 years (SD)) accepted Bangerter filter patching therapy (whole walking time in each day) after a 2-month of refractive adaptation. The ocular dominance index difference was quantified using a binocular rivalry task before the start of the patching therapy. The dashed line is a linear fit of the data. A 2-tailed Pearson correlation analysis showed that the correlation between the amblyopic eye acuity improvement after 2 months of treatment and the ocular dominance index difference was not significant: r = -0.22, *P* = 0.54.

## Discussion

We show that there was no significant correlation between the amblyopic eye acuity improvement after 2 months of occlusion treatment and the normalized ocular dominance (OD) index difference associated with the OD plasticity resulting from the amblyopic eye being subjected to 2 hrs of short-term deprivation.

On the face of it, our results seem to be in consistent with the claim made by Lunghi et al (2016) (Lunghi et al., 2016) that the ocular dominance change measured 2 hours after occlusion therapy predicts “the recovery rate” of the amblyopic eye in anisometropic children. However, Lunghi et al (Lunghi et al., 2016) in their original paper plotted the change in ocular dominance (OD) against the *absolute* acuity at end of treatment. This is because they defined the effects of occlusion treatment (the “recovery rate”) in terms of the absolute visual acuity of the amblyopic eye measured after 2 months of treatment. We believe that this prediction is not interesting because clinically, in patients with amblyopia, their final visual acuity is best predicted by their initial visual acuity before. This is also true in Lunghi et al’s study. According to Table 1 in Lunghi et al’s paper, there is a strong correlation between the amblyopic eye’s visual acuity before and after 2-month of treatment: r = 0.731, *P* = 0.016. The p-value is even smaller than that reported in Lunghi et al’s paper using the homeostatic plasticity (rho = -0.65, *P* = 0.04). This suggests that if one’s objective was to predict amblyopic eye’s visual acuity after 2-month of occlusion therapy, one can simply rely on patients’ initial visual acuity rather than a complicated psychophysics measure. What one really wants to do is to predict what change will occur in acuity as a result of treatment. That is what we set out to do.

Thus, our purpose was to investigate the relationship between the effects of the occlusion therapy, in terms of the visual acuity *improvement (i*.*e*., *the acuity benefit)* of the amblyopic eye measured after 2 months of treatment, and the short-term monocular deprivation induced visual plasticity. Therefore, we plotted *the change* in ocular dominance from short-term deprivation against *the change* in acuity from classical patching, as this is the only valid way of assessing whether changes in short term plasticity can predict improvements in long term patching. According to Amblyopia PPP(Wallace et al., 2018), success of patching therapy only make sense if it is measured in terms of an incremental change, that is an “improvement”. After re-plotting Lunghi et al (2016)’s data in this more convention way (corresponding to our data Figure 3A) where OD changes are plotted against patching-induced acuity changes, their results agree with ours and show that there is no significant correlation (Lunghi et al: r = -0.22, *P* = 0.54; the present study: r = 0.20, *P* = 0.66) between OD changes at the beginning of patching therapy and acuity changes after 2 months of classical patching therapy.

There were a number of important differences between our study and the previous one by Lunghi et al (2016) (Lunghi et al., 2016). However, since the data from the two studies are consistent with there being no significant correlation between short-term ocular dominance plasticity changes and longer-term patching therapy improvements, none of these differences can be particularly crucial. Thus, our conclusions are robust in the face of the differences which are now listed:

First, Lunghi et al (2016) (Lunghi et al., 2016) used binocular rivalry to measure ocular dominance whereas we used a binocular combination task. Both tasks are laboratory-based tests and are potentially hard for children. In Lunghi et al (2016) (Lunghi et al., 2016), they added features to make the test child-friendly. In our study, we instigated practice sessions and only chose patients who were able to do the test accurately such that their R-square values for the “binocularly perceived phase vs. interocular contrast ratio” curve were larger than 0.85. Nevertheless, both tests have been widely used in studying the homeostatic plasticity in normal adults (rivalry:(Lunghi et al., 2011; Lunghi et al., 2013; Lunghi et al., 2015b; Lunghi and Sale, 2015; Bai et al., 2017; Kim et al., 2017; Ramamurthy and Blaser, 2018; Finn et al., 2019; Sheynin et al., 2019a; Sheynin et al., 2019b); combination:(Zhou et al., 2013a; Zhou et al., 2014; Bai et al., 2017; Wang et al., 2017; Yao et al., 2017; Zhou et al., 2017a; Zhou et al., 2017b; Min et al., 2018; Min et al., 2019; Sheynin et al., 2019a)) and in patients with amblyopia (rivalry:(Lunghi et al., 2019); combination:(Zhou et al., 2013c; Zhou et al., 2019)).

Second, Lunghi et al (2016) (Lunghi et al., 2016) measured the ocular dominanceassociated with short-term occlusion of the fellow fixing eye whereas we measured the ocular dominance changes associated with the short-term deprivation of the amblyopic eye. To our limited knowledge, it is so far not clear whether the effect of short-term patching differ in magnitude for patching different eyes. We have no reason to believe that the underlying mechanisms in patching different eyes are different. We are at present assuming this. In support of this assumption, both studies found similar directions of ocular dominance shift (in favor of the patched eye) in most of the patients (9/10 in Lunghi et al’s study and 5/7 in the present study) after the short-term monocular deprivation. Limited by the small sample in the present study, the normalized ocular dominance indices for the 7 patients were not statistically larger than 0 (t (6) = 1.33, *P* = 0.23). However, we still failed to find any significantly correlation between the amblyopic eye acuity improvement after 2 months of occlusion treatment and the normalized ocular dominance (OD) index difference associated with the OD plasticity based on those having an ocular dominance shift in favor of the patched eye (n = 5; r = -0.14, *P* = 0.82).

Third, their patients had mild-moderate amblyopia (<= 0.4 logMAR) and were treated by occluding the fellow eye with a Bangerter filter (strength 0.4) whereas our patients had more severe amblyopic > 0.45 logMAR and were treated by occluding the fellow eye with an opaque patch.

Fourth, the main conclusions in both the Lunghi et al’s study (Lunghi et al., 2016)) and the current study relied on correlations in small samples (i.e., 7 in ours and 10 in Lunghi et al (2016) (Lunghi et al., 2016)). It’s always hard to justify what’s the proper sample size for a valid conclusion based on a correlation analysis, e.g., why 10 is enough, while 7 is not acceptable? This itself is tightly linked to the question that one asks. In particular, for the question that we asked, whether the homeostatic plasticity predicts the recovery rate (or the effects of occlusion therapy; or the visual acuity improvement), both Lunghi et al’s and our study failed to reach a significant correlation. Thus, both of these two studies suggest that the homeostatic plasticity might not be able to predict the acuity improvements from occlusion therapy. Considering that this conclusion relies on 2 studies (Lunghi et al’s and ours) with 17 patients (10 in Lunghi et al’s and 7 in ours) from 2 independent groups using different techniques, we believe that this strengthens the conclusion. In other words, if one has to get a large sample to reach significance in this kind of correlation analysis, it’s hard to believe that we can use the homeostatic plasticity as a prediction index in clinical practice.

Short-term ocular dominance plasticity does not provide an index of cortical plasticity in the general sense, such that it could be used to predict acuity improvement outcomes from classical patching. This conclusion is robust to the type of measurement method used, the degree of amblyopia treated and the eye that is occluded (i.e., fixing vs amblyopic) in the short-term OD measurement. In some ways this general conclusion is unsurprising because short term OD plasticity is a very specific homeostatic form of plasticity(Min et al., 2018), which may be quite different from the type of plasticity that underlies monocular patching therapy. While it may not predict monocular outcomes in patching therapy, it is yet to be determined if this is also true for purely binocular outcomes, using purely binocular therapy (Hess and Thompson, 2015), or changes of neural transmitter concentration (e.g. GABA(Lunghi et al., 2015b)), etc.

All that can be said is that there appears to be no simple straightforward prediction from the extent of ocular dominance change from short-term occlusion and the acuity benefit of long-term occlusion therapy.

## Data Availability

The datasets generated for this study are available on request to
the corresponding authors.

## DATA AVAILABILITY

The datasets generated for this study are available on request to the corresponding author.

## ETHICS STATEMENT

This study was carried out in accordance with the recommendations of the ethics committee of the Wenzhou Medical University, with written informed consent from all subjects after explanation of the nature and possible consequences of the study. All subjects gave written informed consent in accordance with the Declaration of Helsinki. The protocol was approved by the ethics committee of the Institutional Review Boards of Wenzhou Medical University.

## AUTHOR CONTRIBUTIONS

CT, JZ, and RH conceived the experiments. CT, ZH, and YC performed the experiments. CT, ZH, YC, and JZ analyzed the data and interpreted the data. CT, JZ, and RH wrote the manuscript. All authors contributed to manuscript revision, read and approved the submitted version.

